# A Novel Easy-To-Access Model That Predicts Stroke-Associated Pneumonia

**DOI:** 10.1101/2023.07.13.23292637

**Authors:** Cheng-Yu Chung, Wen-Hwa Wang, Chun-Hao Yin, Jin-Shuen Chen, Yao-Shen Chen, Cheng-Chang Yen, Ching-Huang Lin

**Author notes:** Corresponding Author: Ching-Huang Lin.

## Abstract

**BACKGROUND:** Stroke-associated pneumonia (SAP) is a common poststroke complication but the influence of early neurological deterioration (END) on SAP risk remains unclear. We aimed to develop an easy-to-access model to predict SAP and evaluate the SAP–END relationship using the Glasgow Coma Scale (GCS).

**METHODS:** This retrospective study enrolled consecutive patients aged ≥20 years with first-ever acute ischemic stroke at Kaohsiung Veterans General Hospital between January 1, 2010, and November 30, 2020. SAP was defined according to modified Centers for Disease Control and Prevention criteria. Patients’ baseline characteristics, laboratory data within 24 h, neurological findings, and serial GCS scores within 48 h were collected. Regression analysis was used to identify independent risk factors for SAP.

**RESULTS:** Among 1009 enrolled patients, SAP occurred in 85 (8.4%) patients. Using multivariate analysis, END after admission (adjusted odds ratio [aOR] 2.94, 95% confidence interval [CI] 1.74–4.95, P<0.001) remained significant after adjusting for confounders. Initial GCS score <10 (aOR 2.30, 95% CI 1.30–4.06, P=0.004), National Institutes of Health Stroke Scale (NIHSS) score 5–15 (aOR 2.02, 95% CI 1.09–3.73, P=0.026) or ≥16 (aOR 3.45, 95% CI 1.72–6.89, P<0.001), cardioembolism (aOR 4.76, 95% CI 1.90–11.91, P=0.014), undetermined etiology (aOR 3.41, 95% CI 1.29–9.03, P=0.001), and neutrophil-to-lymphocyte ratio (NLR) >2.5 (aOR 2.10, 95% CI 1.28–3.46, P=0.004) were also significant. The area under the curve (AUC) of combined GCS score, END, NIHSS score, cardioembolism, stroke with undetermined type, and NLR was 0.83 (95% CI 0.78–0.87, P<0.001), which was superior to that of PANTHERIS scores (0.62, 95% CI 0.55–0.68, P<0.001).

**CONCLUSIONS:** This study developed a simple predictive model for SAP using easily accessible and generally available parameters. GCS-based END was an independent risk factor for SAP. Early identification of SAP risk factors and reversible END causes may lower SAP incidence.

## INTRODUCTION

Stroke-associated pneumonia (SAP), a common stroke complication, affects, depending on SAP definitions and patient characteristics, 5.6–37.98% of patients with acute ischemic stroke.^1–7^ Patients with SAP are affected by longer lengths of hospital stay, more expensive hospitalization, higher mortality, and poor long-term prognosis compared to those without.^6^

Diagnosing SAP is challenging, and the diagnostic sensitivity of chest radiography is low in the early disease stages.^8^ Several risk factors have been identified, including age, smoking, initial stroke severity, impaired consciousness, dysphagia, congestive heart failure, atrial fibrillation, prestroke dependence, hyperglycemia, and indwelling nasogastric tubes, and several predictor scores have been developed.^1–5, 7^ However, the utility of such scores in routine clinical care remains unclear, and some items such as dysphagia screening not only require sophisticated evaluation but also lack standardization, making SAP prediction complex.

To predict SAP as early as possible, most models focus on baseline clinical features. Substantial early neurological deterioration (END) may occur in the acute phase of ischemic stroke, causing SAP underestimation by these scores. END, also known as “stroke-in-evolution” or “progressing stroke,” has been observed in 11.8–43% of patients with stroke, depending on the definition used.^9–14^ END is commonly defined as a 2-point decrease on the Glasgow Coma Scale (GCS) or a change of at least 1 point on the Canadian Neurological Scale or at least 2 points on the Scandinavian Stroke Scale or National Institutes of Health Stroke Scale (NIHSS). As no international consensus exists, different definitions have been used in recent studies. END is associated with poor long-term outcomes,^11–14^ but little is known about the association between END and complications in the acute phase of stroke, especially SAP.

Here, we aimed to develop a novel and easily accessible model to predict SAP and evaluate the relationship between END and SAP in patients with acute stroke.

## METHODS

### Patient Enrollment

This retrospective study was conducted at Kaohsiung Veterans General Hospital (KVGH), a 1700-bed medical center providing primary and tertiary referral care in southern Taiwan.

To identify patients with ischemic stroke, we used the 9th revision of the International Classification of Diseases (ICD-9: 434, 436, 437.1) and the 10th revision of the International Statistical Classification of Diseases and Related Health Problems (ICD-10: I63, I65, I66, and I67) with primary, first secondary, second secondary, or third secondary diagnosis codes.^15, 16^

To define “acute stroke,” we combined ICD codes with the 30-day “catastrophic illness certificate.” In Taiwan, patients with severe illnesses, including acute ischemic stroke, can apply for this temporary certificate up to 30 days after stroke onset. Patients receiving care for the illness or related conditions within the certificate’s validity period do not need to make a copayment for outpatient or inpatient care. All applications for catastrophic illness certificates are reviewed by experts.^17^

Consecutive hospitalized patients with first-ever acute ischemic stroke between January 1, 2010, and November 30, 2020, aged ≥20 years, with thorough stroke severity evaluation using the NIHSS at hospital arrival, and serial GCS data (at least recorded at hospital arrival, 24 h after admission, and 48 h after admission) were included. Patients with an active infection at admission, incomplete laboratory data, or pneumonia developing within 24 h of hospital arrival were excluded. Finally, 1009 patients were enrolled (Figure 1) with ICD-10: I63 accounting for the majority (n=962, 95.3%) and ICD-9: 434, ICD-10: I65, I66, and I67 for the rest (n=47, 4.7%).

**Figure 1.**
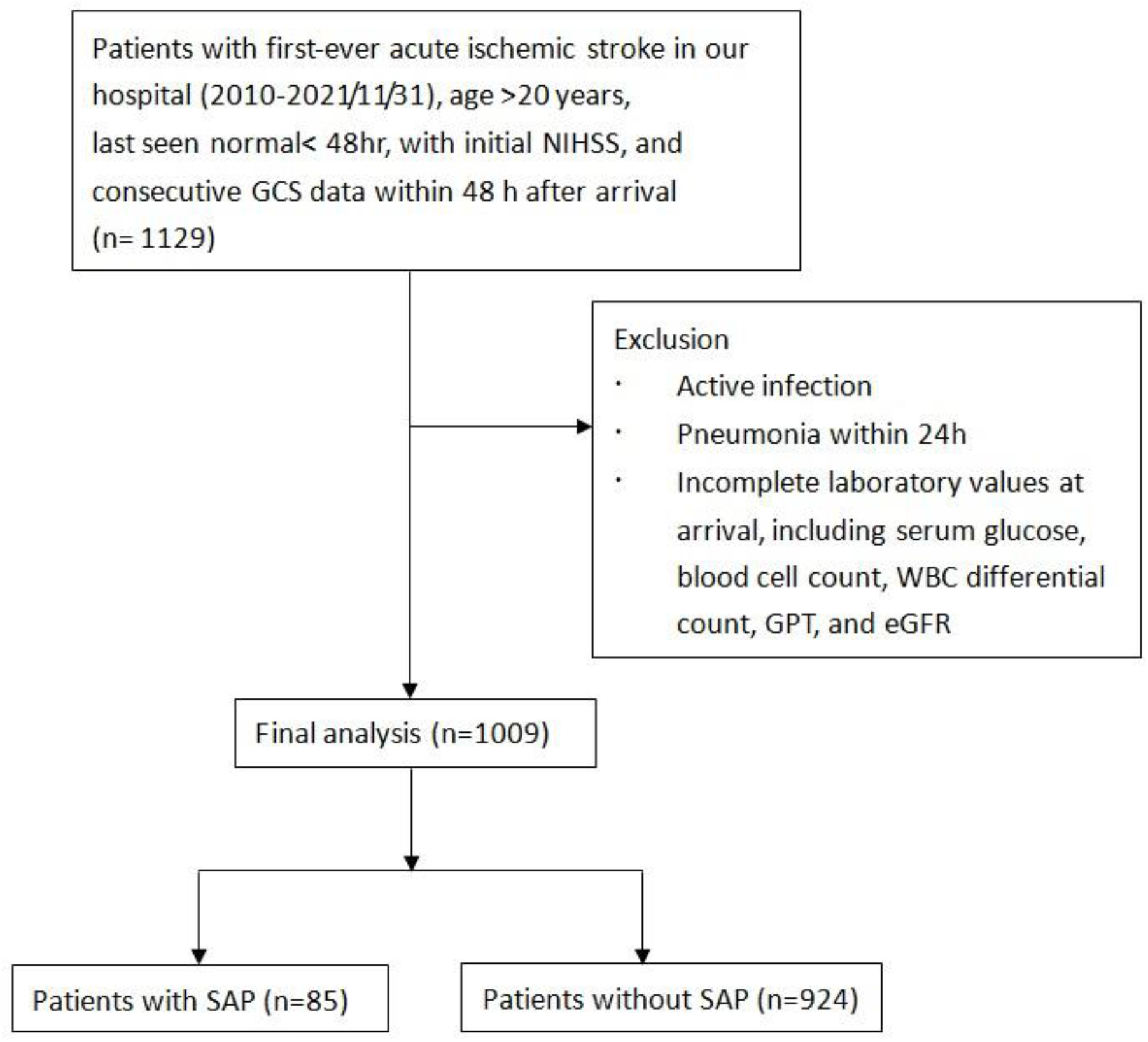
Flowchart of patient enrolment. eGFR indicates estimated glomerular filtration rate; GPT, glutamic pyruvic transaminase; GCS, Glasgow Coma Scale; NIHSS, National Institute of Health Stroke Scale; SAP, stroke- associated pneumonia; and WBC, white blood cell.

### Definitions

#### SAP Diagnosis

The primary outcome was SAP referring to lower respiratory tract infections that occur within the first 7 days after stroke onset. According to modified Centers for Disease Control and Prevention criteria, SAP can be classified as “probable SAP” and “definite SAP.”^8^ In the present study, the diagnosis of SAP was based on the discharge diagnosis reported by the treating physician according to clinical symptoms, physical examination, laboratory data, culture results, or radiological findings. Combined with the onset, the diagnosis fitted at least in the category “probable SAP.”

SAP onset was defined as the timing of empiric antibiotic therapy reflected by Anatomical Therapeutic Chemical codes, including those for amoxicillin with/without clavulanic acid, ampicillin with/without sulbactam, piperacillin with/without tazobactam, cefoxitin, ceftriaxone, cefoperazone/sulbactam, levofloxacin, moxifloxacin, and ciprofloxacin.

#### Early Neurological Deterioration

END was defined as a decrease of ≥2 on the GCS between serial evaluations in the first 48 h of hospitalization with an interval of 24 h.

#### TOAST Classification

Stroke subtypes were classified according to the Trial of Org 10172 Acute Stroke Treatment (TOAST) classification.^18^

### Data Collection

The following demographic data of each patient were collected from the hospital medical records: age, sex, alcohol consumption, smoking status, underlying comorbidities by ICD-9 and ICD-10, including atrial fibrillation, chronic obstructive pulmonary disease, coronary artery disease, heart failure, diabetes mellitus, hypertension, renal failure, blood pressure (for the PANTHERIS score),^1^ laboratory data obtained within 24 h after hospital arrival, daily GCS assessment, and clinical features of stroke including initial stroke severity according to the NIHSS score, stroke subtype according to TOAST classification, infarction site, administration of tissue plasminogen activator, and endovascular thrombectomy.

### Statistical Analysis

The baseline demographic data of the patients were compared between the groups with and without SAP. Chi-square or Fisher’s exact tests were used to compare categorical data expressed as numbers (percentages), when appropriate. Continuous variables with normal distribution were analyzed as means and standard deviations, and the independent Student’s t-test was used to compare these continuous variables. Univariate and stepwise logistic regression analyses were used to examine independent predictors of SAP occurrence.

Stepwise regression within the backward selection process began with a univariate analysis of the parameters. Variables with significant univariate tests (P<0.1) were selected as candidates for multivariate analyses.^19^ The maximum value of the area under the curve (AUC) was selected to determine the optimal cutoff value for each parameter. Calibration curves were plotted to assess the regression, along with the Hosmer–Lemeshow test. A significant test statistic implies that the model is not perfectly calibrated. The area under the receiver operating characteristic curve was used to evaluate the performance of multivariate logistic regression models. Significance was defined as P<0.05 for all two-tailed tests. All statistical analyses were performed using SAS (version 9.4; SAS System for Windows) and SPSS for Windows (version 20.0; SPSS Inc., Chicago, IL, USA).

### Ethics Approval and Patient Consent

The study was approved by the ethics committee of KVGH (KSVGH22-CT4-12). The requirement for patient consent was waived because of the retrospective observational study design and because the study involved no increase in health risks to any patient.

### Data Availability Statement

Anonymized data will be made available upon reasonable request from qualified investigators.

## RESULTS

Overall, 1009 patients were finally evaluated, with a mean age of 69.1±13.8 years, including 613 male (61%) and 396 female (39%) patients. The mean initial NIHSS score in the cohort was 6.7±8.8. Among these patients, 85 (8.4%) developed SAP. SAP occurred within the first 7 days, mostly (55/85, 62.4%) within the first 2–3 days. Patients with SAP had higher in-hospital mortality than those without (23.5% vs. 3.0%, P<0.001).

Baseline characteristics of the groups with and without SAP are shown in Tables 1 and 2. The patients in the SAP group were older and had higher rates of atrial fibrillation, coronary artery disease, and heart failure than those in the other group. Moreover, serum glucose levels, white blood cell counts, and neutrophil-to-lymphocyte ratios (NLRs) were significantly higher in the group with than in the group without SAP. Furthermore, the estimated glomerular filtration rates and platelet counts were significantly lower in the SAP group. The SAP group also had more severe stroke cases, a greater number of patients with END, higher rates of hemorrhagic transformation, poststroke seizure, and cardioembolism but fewer small-vessel diseases than the group without SAP.

**Table 1.**
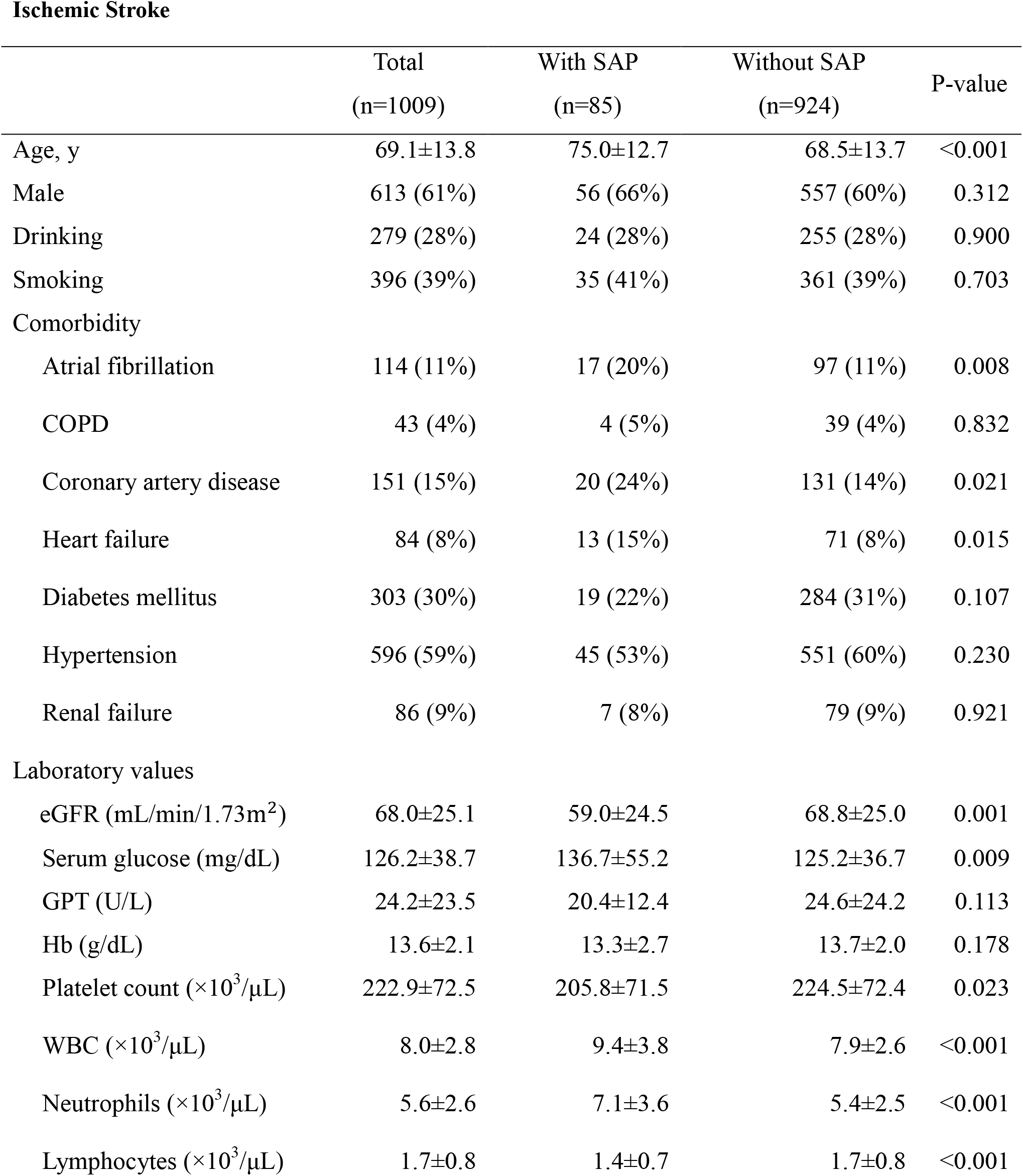
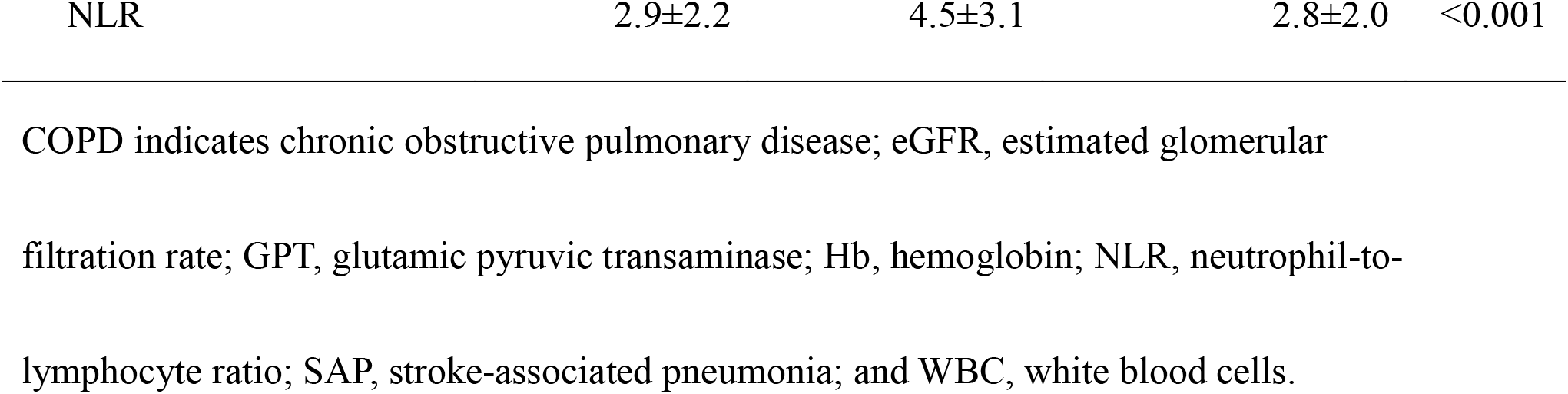
Baseline Characteristics and Laboratory Data at Arrival in Patients With Acute Ischemic Stroke.

Ischemic Stroke
|  | Total<br>(n=1009) | With SAP<br>(n=85) | Without SAP<br>(n=924) | P-value |
| --- | --- | --- | --- | --- |
| Age, y | 69.1±13.8 | 75.0±12.7 | 68.5±13.7 | <0.001 |
| Male | 613 (61%) | 56 (66%) | 557 (60%) | 0.312 |
| Drinking | 279 (28%) | 24 (28%) | 255 (28%) | 0.900 |
| Smoking | 396 (39%) | 35 (41%) | 361 (39%) | 0.703 |
| Comorbidity |  |  |  |  |
| Atrial fibrillation | 114 (11%) | 17 (20%) | 97 (11%) | 0.008 |
| COPD | 43 (4%) | 4 (5%) | 39 (4%) | 0.832 |
| Coronary artery disease | 151 (15%) | 20 (24%) | 131 (14%) | 0.021 |
| Heart failure | 84 (8%) | 13 (15%) | 71 (8%) | 0.015 |
| Diabetes mellitus | 303 (30%) | 19 (22%) | 284 (31%) | 0.107 |
| Hypertension | 596 (59%) | 45 (53%) | 551 (60%) | 0.230 |
| Renal failure | 86 (9%) | 7 (8%) | 79 (9%) | 0.921 |
| Laboratory values |  |  |  |  |
| eGFR (mL/min/1.73m<sup>2</sup>) | 68.0±25.1 | 59.0±24.5 | 68.8±25.0 | 0.001 |
| Serum glucose (mg/dL) | 126.2±38.7 | 136.7±55.2 | 125.2±36.7 | 0.009 |
| GPT (U/L) | 24.2±23.5 | 20.4±12.4 | 24.6±24.2 | 0.113 |
| Hb (g/dL) | 13.6±2.1 | 13.3±2.7 | 13.7±2.0 | 0.178 |
| Platelet count (×10<sup>3</sup>/μL) | 222.9±72.5 | 205.8±71.5 | 224.5±72.4 | 0.023 |
| WBC (×10<sup>3</sup>/μL) | 8.0±2.8 | 9.4±3.8 | 7.9±2.6 | <0.001 |
| Neutrophils (×10<sup>3</sup>/μL) | 5.6±2.6 | 7.1±3.6 | 5.4±2.5 | <0.001 |
| Lymphocytes (×10<sup>3</sup>/μL) | 1.7±0.8 | 1.4±0.7 | 1.7±0.8 | <0.001 |
| NLR | 2.9±2.2 | 4.5±3.1 | 2.8±2.0 | <0.001 |

COPD indicates chronic obstructive pulmonary disease; eGFR, estimated glomerular filtration rate; GPT, glutamic pyruvic transaminase; Hb, hemoglobin; NLR, neutrophil-to-lymphocyte ratio; SAP, stroke-associated pneumonia; and WBC, white blood cells.

**Table 2.**
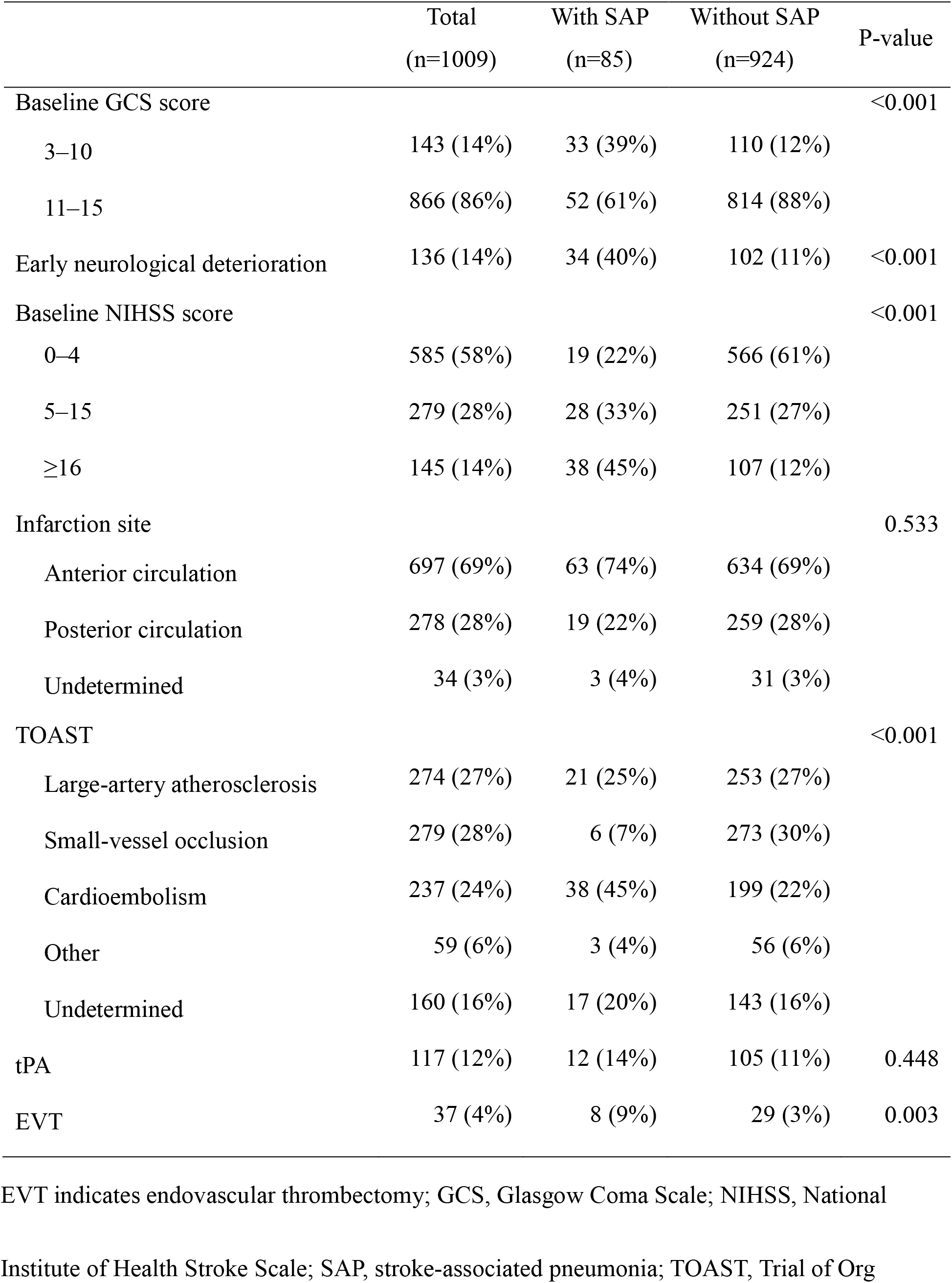
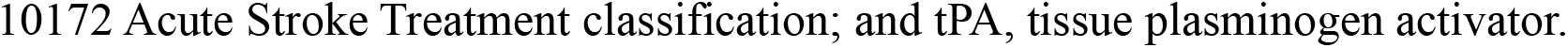
Neurologic Characteristics of Patients With Acute Ischemic Stroke.

|  | Total<br>(n=1009) | With SAP<br>(n=85) | Without SAP<br>(n=924) | P-value |
| --- | --- | --- | --- | --- |
| Baseline GCS score |  |  |  | <0.001 |
| 3–10 | 143 (14%) | 33 (39%) | 110 (12%) |  |
| 11–15 | 866 (86%) | 52 (61%) | 814 (88%) |  |
| Early neurological deterioration | 136 (14%) | 34 (40%) | 102 (11%) | <0.001 |
| Baseline NIHSS score |  |  |  | <0.001 |
| 0–4 | 585 (58%) | 19 (22%) | 566 (61%) |  |
| 5–15 | 279 (28%) | 28 (33%) | 251 (27%) |  |
| ≥16 | 145 (14%) | 38 (45%) | 107 (12%) |  |
| Infarction site |  |  |  | 0.533 |
| Anterior circulation | 697 (69%) | 63 (74%) | 634 (69%) |  |
| Posterior circulation | 278 (28%) | 19 (22%) | 259 (28%) |  |
| Undetermined | 34 (3%) | 3 (4%) | 31 (3%) |  |
| TOAST |  |  |  | <0.001 |
| Large-artery atherosclerosis | 274 (27%) | 21 (25%) | 253 (27%) |  |
| Small-vessel occlusion | 279 (28%) | 6 (7%) | 273 (30%) |  |
| Cardioembolism | 237 (24%) | 38 (45%) | 199 (22%) |  |
| Other | 59 (6%) | 3 (4%) | 56 (6%) |  |
| Undetermined | 160 (16%) | 17 (20%) | 143 (16%) |  |
| tPA | 117 (12%) | 12 (14%) | 105 (11%) | 0.448 |
| EVT | 37 (4%) | 8 (9%) | 29 (3%) | 0.003 |

EVT indicates endovascular thrombectomy; GCS, Glasgow Coma Scale; NIHSS, National
Institute of Health Stroke Scale; SAP, stroke-associated pneumonia; TOAST, Trial of Org
10172 Acute Stroke Treatment classification; and tPA, tissue plasminogen activator.

The associations between baseline data at hospital arrival, stroke features, and SAP incidence in the univariate analysis are shown in Table 3. The following variables with P<0.1 in the univariate analysis were included in the multivariate analysis: age >65 years, atrial fibrillation, coronary artery disease, heart failure, estimated glomerular filtration rate <30 mL/min/1.73, serum glucose >200 mg/dL, platelet count <200×10^3^/μL NLR >2.5, initial NIHSS score (5–15 and ≥16), baseline GCS score <10, neurological deterioration, cardioembolism, and stroke of undetermined etiology.

**Table 3.**
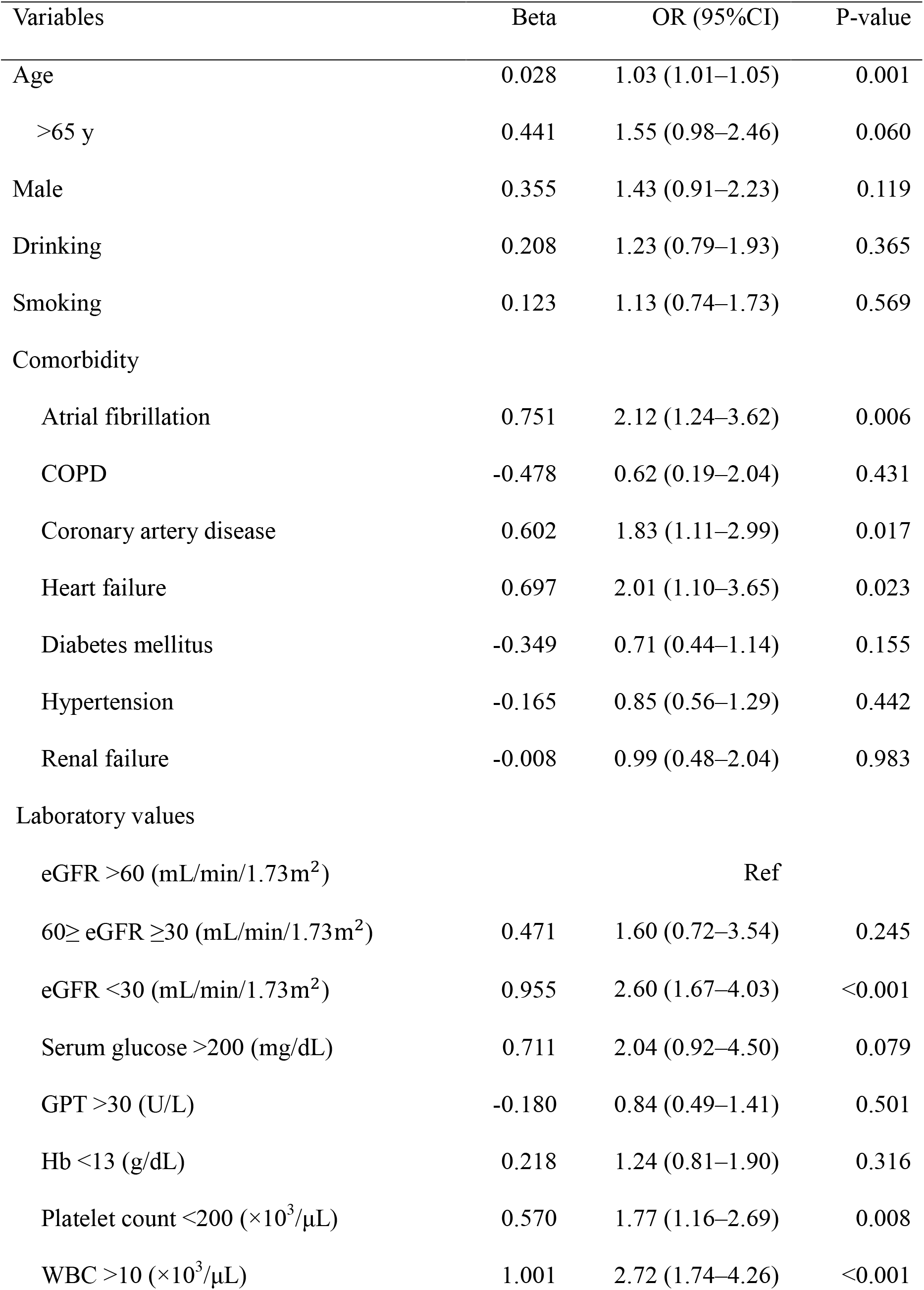
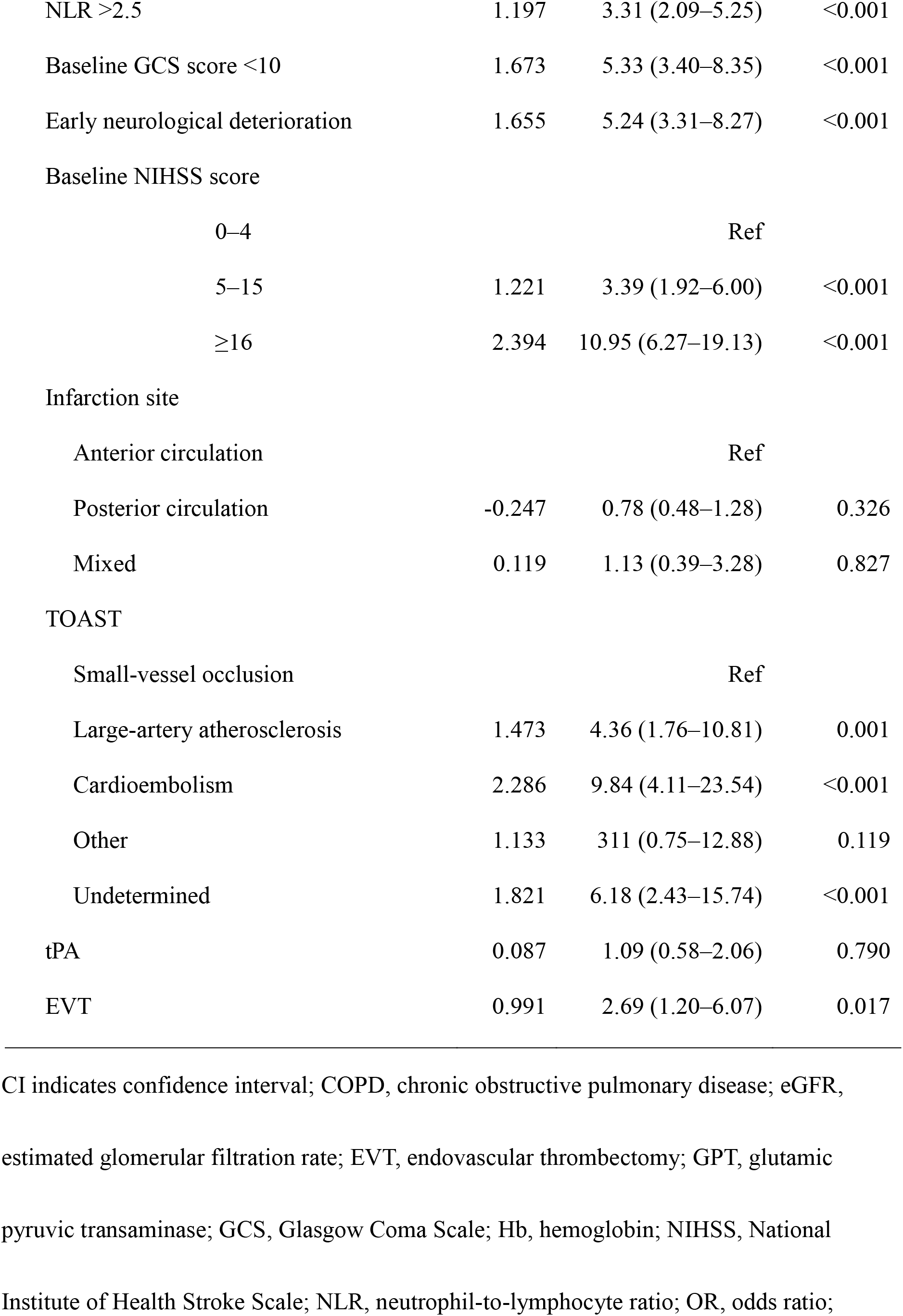
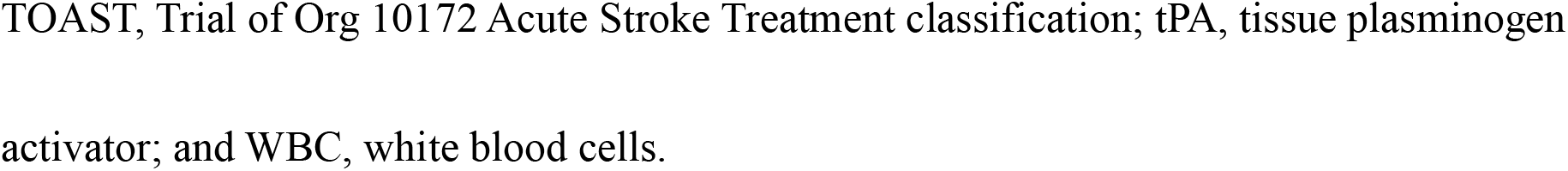
Univariate Logistic Regression Analysis for Stroke-Associated Pneumonia.

| Variables | Beta | OR (95%CI) | P-value |
| --- | --- | --- | --- |
| Age | 0.028 | 1.03 (1.01–1.05) | 0.001 |
| >65 y | 0.441 | 1.55 (0.98–2.46) | 0.060 |
| Male | 0.355 | 1.43 (0.91–2.23) | 0.119 |
| Drinking | 0.208 | 1.23 (0.79–1.93) | 0.365 |
| Smoking | 0.123 | 1.13 (0.74–1.73) | 0.569 |
| Comorbidity |  |  |  |
| Atrial fibrillation | 0.751 | 2.12 (1.24–3.62) | 0.006 |
| COPD | -0.478 | 0.62 (0.19–2.04) | 0.431 |
| Coronary artery disease | 0.602 | 1.83 (1.11–2.99) | 0.017 |
| Heart failure | 0.697 | 2.01 (1.10–3.65) | 0.023 |
| Diabetes mellitus | -0.349 | 0.71 (0.44–1.14) | 0.155 |
| Hypertension | -0.165 | 0.85 (0.56–1.29) | 0.442 |
| Renal failure | -0.008 | 0.99 (0.48–2.04) | 0.983 |
| Laboratory values |  |  |  |
| eGFR >60 (mL/min/1.73m<sup>2</sup>) |  | Ref |  |
| 60≥ eGFR ≥30 (mL/min/1.73m<sup>2</sup>) | 0.471 | 1.60 (0.72–3.54) | 0.245 |
| eGFR <30 (mL/min/1.73m<sup>2</sup>) | 0.955 | 2.60 (1.67–4.03) | <0.001 |
| Serum glucose >200 (mg/dL) | 0.711 | 2.04 (0.92–4.50) | 0.079 |
| GPT >30 (U/L) | -0.180 | 0.84 (0.49–1.41) | 0.501 |
| Hb <13 (g/dL) | 0.218 | 1.24 (0.81–1.90) | 0.316 |
| Platelet count <200 (×10<sup>3</sup>/μL) | 0.570 | 1.77 (1.16–2.69) | 0.008 |
| WBC >10 (×10<sup>3</sup>/μL) | 1.001 | 2.72 (1.74–4.26) | <0.001 |
| NLR >2.5 | 1.197 | 3.31 (2.09–5.25) | <0.001 |
| Baseline GCS score <10 | 1.673 | 5.33 (3.40–8.35) | <0.001 |
| Early neurological deterioration | 1.655 | 5.24 (3.31–8.27) | <0.001 |
| Baseline NIHSS score |  |  |  |
| 0–4 |  | Ref |  |
| 5–15 | 1.221 | 3.39 (1.92–6.00) | <0.001 |
| ≥16 | 2.394 | 10.95 (6.27–19.13) | <0.001 |
| Infarction site |  |  |  |
| Anterior circulation |  | Ref |  |
| Posterior circulation | -0.247 | 0.78 (0.48–1.28) | 0.326 |
| Mixed | 0.119 | 1.13 (0.39–3.28) | 0.827 |
| TOAST |  |  |  |
| Small-vessel occlusion |  | Ref |  |
| Large-artery atherosclerosis | 1.473 | 4.36 (1.76–10.81) | 0.001 |
| Cardioembolism | 2.286 | 9.84 (4.11–23.54) | <0.001 |
| Other | 1.133 | 3.11 (0.75–12.88) | 0.119 |
| Undetermined | 1.821 | 6.18 (2.43–15.74) | <0.001 |
| tPA | 0.087 | 1.09 (0.58–2.06) | 0.790 |
| EVT | 0.991 | 2.69 (1.20–6.07) | 0.017 |

CI indicates confidence interval; COPD, chronic obstructive pulmonary disease; eGFR, estimated glomerular filtration rate; EVT, endovascular thrombectomy; GPT, glutamic pyruvic transaminase; GCS, Glasgow Coma Scale; Hb, hemoglobin; NIHSS, National Institute of Health Stroke Scale; NLR, neutrophil-to-lymphocyte ratio; OR, odds ratio;
TOAST, Trial of Org 10172 Acute Stroke Treatment classification; tPA, tissue plasminogen activator; and WBC, white blood cells.

### Multivariate Logistic Regression Analysis of SAP Risk Factors

Results of the multivariate analysis revealed that baseline GCS score (<10: odds ratio [OR] 2.30, 95% confidence interval [CI] 1.30–4.06, P=0.004), NIHSS score (5–15: OR 2.02, 95% CI 1.09–3.73, P=0.026; ≥16: OR 3.45, 95% CI 1.72–6.89, P<0.001), neurological deterioration after arrival (OR 2.94, 95% CI 1.74–4.95, P<0.001), NLR (OR 2.10, 95% CI 1.28–3.46, P=0.004), and stroke subtype (cardioembolism: OR 4.76, 95% CI 1.90–11.91, P=0.001; undetermined etiology: OR 3.41, 95% CI 1.29–9.03, P=0.014) were independent risk factors for SAP (Table 4).

**Table 4.** Stepwise Logistic Regression Analysis for Risk of Stroke-Associated Pneumonia.

| Variables | Beta | aOR (95% CI) | P-value |
| --- | --- | --- | --- |
| Baseline GCS score <10 | 0.833 | 2.30 (1.30–4.06) | 0.004 |
| Early neurological deterioration | 1.077 | 2.94 (1.74–4.95) | <0.001 |
| Baseline NIHSS score |  |  |  |
| 0–4 |  | Ref |  |
| 5–15 | 0.700 | 2.02 (1.09–3.73) | 0.026 |
| ≥16 | 1.237 | 3.45 (1.72–6.89) | <0.001 |
| NLR >2.5 | 0.743 | 2.10 (1.28–3.46) | 0.004 |
| TOAST |  |  |  |
| Small-vessel occlusion |  | Ref |  |
| Large-artery atherosclerosis | 0.877 | 2.40 (0.96–6.19) | 0.069 |
| Cardioembolism | 1.559 | 4.76 (1.90–11.91) | 0.001 |
| Other | 0.524 | 1.69 (0.39–7.34) | 0.485 |
| Undetermined | 1.226 | 3.41 (1.29–9.03) | 0.014 |
| c-statistic |  | 0.825 (0.782–0.868) | <0.001 |
| Hosmer–Lemeshow test |  | Chi-square=3.794 | 0.803 |

aOR indicates adjusted odds ratio; CI, confidence interval; GCS, Glasgow Coma Scale;
TOAST, Trial of Org 10172 Acute Stroke Treatment classification.

### Predictive Effect of Identified Risk Factors

Receiver operating characteristic curve analysis showed that the AUC of the identified risk factors in predicting SAP was 0.83 (95% CI 0.78–0.87, P<0.001), which was in our population superior to the PANTHERIS score AUC of 0.62 (95% CI 0.55–0.68, P<0.001; Figure 2).

**Figure 2.**
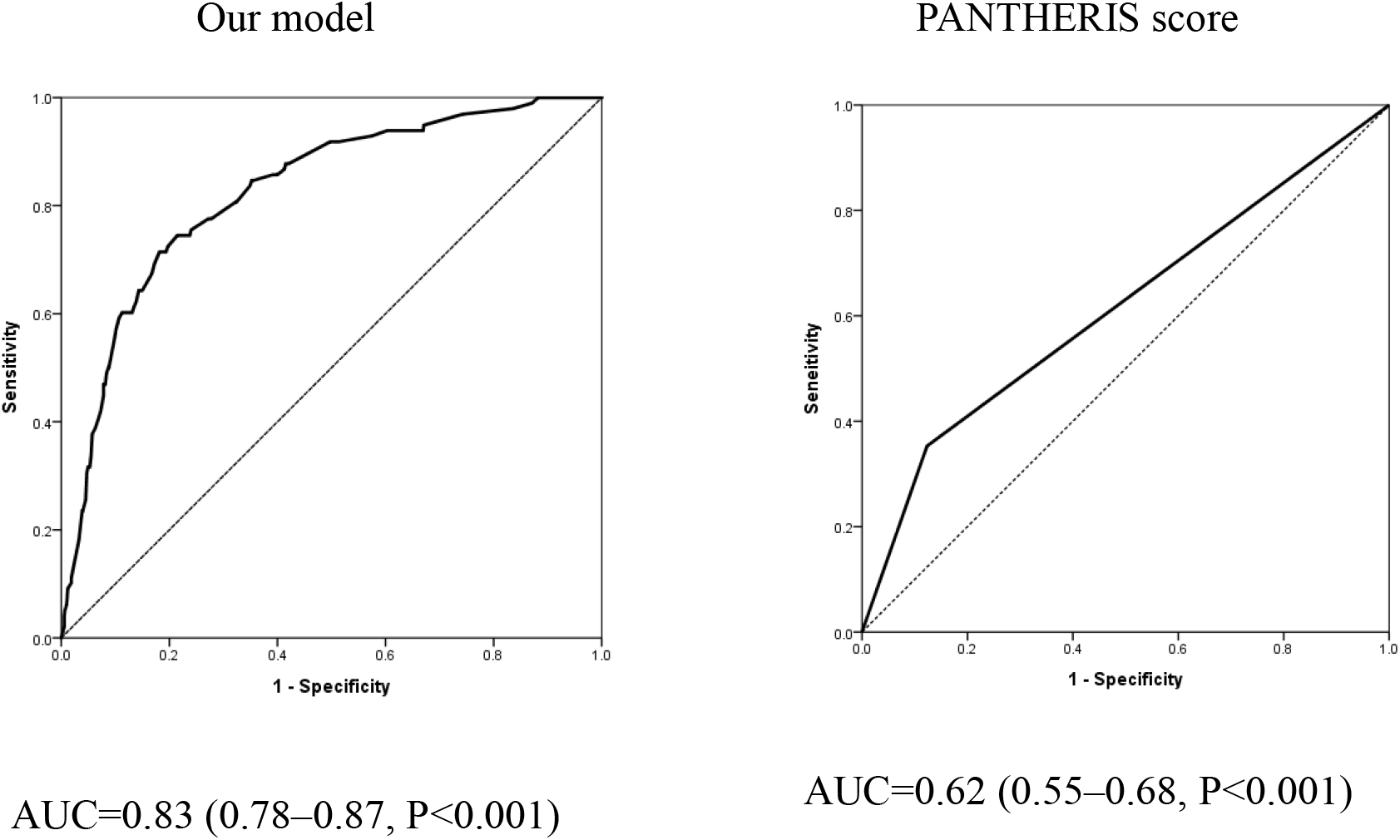
Comparison of area under the receiver operating characteristic curve (AUC) values between our model (combined risk factors of GCS and NIHSS scores, early neurological deterioration, NLR, and stroke subtype) and the PANTHERIS score for SAP prediction. GCS indicates Glasgow Coma Scale; NIHSS, National Institute of Health Stroke Scale; NLR, neutrophil-to-lymphocyte ratio; and SAP, stroke-associated pneumonia.

## DISCUSSION

In our cohort, SAP occurred mostly within the first 2–3 days, as previously reported,^20^ and patients with SAP had higher in-hospital mortality than those without. Moreover, SAP has been proven to be a strong risk factor for poor functional outcomes.^4, 6^ Early identification of SAP risk with proper intervention may lower the chances of poor outcomes and decrease medical expenses.

We retrospectively analyzed the risk factors for SAP. Although the impact of END after admission of stroke patients on SAP remains unclear, an association between END and SAP has been reported. In previous studies, half of the END cases occurred within 48 h of the last reported normal state.^13, 21^ This is the main reason why we focused on GCS changes in the first 48 h after hospital arrival.

We found that poor consciousness based on GCS scores, greater stroke severity based on NIHSS scores, higher NLR, and specific stroke subtypes, both cardioembolism and undetermined etiology, were significantly associated with SAP.

Patients with impaired consciousness are at greater risk of aspiration and infectious complications.^22^ Nevertheless, whether patients with END have similar risks regardless of whether they have moderate to severe consciousness disturbance (GCS score<10) is unknown. In this study, 34 patients had both END and SAP, and all had END at or within 2 days before SAP onset. Retrospectively, deteriorating GCS scores occurred before pneumonia was diagnosed. We cannot distinguish whether the altered mental status was a result of neurological complications or an early pneumonia symptom, but among patients with END, those with SAP had a significantly higher proportion of neurological complications (intracranial hemorrhage, seizure, or both) compared to those without SAP (n=14/41, 41% vs. n=19/102, 19%, P=0.008). However, separate etiologies did not significantly differ.

Another study compared the complication rates between patients with and without END, including pneumonia after admission.^13^ They reported higher rates of in-hospital mortality and institutionalization but no significant difference in pneumonia occurrence, possibly because the definitions of END and pneumonia, rather than SAP exclusively, differed from those in our study.

Similar to previous studies, we observed a higher chance of END in patients with greater initial stroke severity,^13^ and this finding persisted after adjusting for NIHSS and GCS scores. A considerable percentage of patients with END presented only relatively mild conscious disturbances (GCS score ≥10) complicated with SAP (16/34, 47.1%), and their risks can be initially overlooked. To support this hypothesis, we applied the GCS-centric 12-point PANTHERIS system and compared its utility to that of our model.^1^ Our model, which included not only baseline neurological features but also GCS changes within 48 h after hospital arrival, turned out to have superior predictive value. Notably, the PANTERIS score was originally developed for patients with severe stroke in the middle cerebral artery territory admitted to neurocritical care units in Germany. In contrast, the patients included in our study were mostly admitted to the emergency or neurology departments. Moreover, the PANTHERIS score was derived from a relatively small single-center cohort, limiting its generalizability.^23^ This may explain its lower predictive value in our population in addition to differences in model items.

Many studies have described potential mechanisms causing END, including collateral failure, clot progression, reocclusion, recurrent stroke, infection, metabolic abnormalities, cerebral edema, hemorrhagic transformation, seizures, hemodynamic factors, excitotoxicity, and inflammatory mechanisms.^24, 25^ However, only a few studies have reported causes of END other than symptomatic intracranial hemorrhage. Even taking the most cited causes into account (malignant edema, early recurrent ischemic stroke, early seizure attack), a large fraction of END seemingly has no immediately identifiable mechanism.^26^ In our study of 1009 patients with first-ever stroke, 136 (13.5%) experienced END during the acute phase.

Among them, identifiable causes of END were hemorrhagic transformation (n=25, 18%), seizure (n=5, 4%), and both (n=3, 2%).

Regarding tools defining END, the NIHSS score, a measure of stroke severity, is commonly used in daily practice, stroke registries, and clinical trials. However, the NIHSS is only valid when used by certified physicians/nurses with appropriate training, rather than by medical staff involved in the primary care of patients with stroke. Heterogeneity in NIHSS scores remained even among certified examiners despite repeated training.^27, 28^ Although the NIHSS is widely used to determine whether patients with acute stroke are eligible for recanalization by tissue plasminogen activator administration or endovascular thrombectomy, the NIHSS was not designed to serve as a bedside rating tool.^29^ To develop a simple prediction model for SAP that can be used by multidisciplinary staff, including resident physicians and nurses in every department, we chose the daily GCS score as a parameter for neurological deterioration.

It has been reported that patients with dysphagia have up to 10-fold increased risks of developing SAP.^2, 3, 30, 31^ Nevertheless, screening methods for dysphagia are diverse and have different diagnostic utilities. According to the Taiwan Clinical Performance Indicator, every patient with suspected acute stroke undergoes dysphagia screening using a bedside water-drinking test on admission. However, screening methods are not standardized, and differences exist among hospitals throughout the country. Previous studies demonstrated that the NIHSS score is correlated with dysphagia and age.^5^ One study reported that for SAP, the NIHSS score had a better predictive value than a dysphagia screening tool.^32^ To make the predictive model easily accessible and enhance its practicality, we did not include dysphagia in our cohort, although it plays a crucial role in SAP.

Atrial fibrillation is a risk factor for SAP in many studies,^2–5^ with several explanations for this relationship. During atrial fibrillation, irregular atrial activities can decrease cardiac output, which causes pulmonary congestion that subsequently makes patients vulnerable to pulmonary infections.^33^ Furthermore, cardioembolic stroke is quite severe, with subsequent cerebral edema and possible neurological deterioration after onset.^34^ Residual confounding for stroke severity cannot be excluded, although this finding was still present after adjustment for NIHSS and GCS scores. Interestingly, among stroke subtypes according to the TOAST classification, cardioembolism, rather than a history or documented atrial fibrillation at admission, appeared to be significantly associated with SAP in this study. In most patients, atrial fibrillation was diagnosed after stroke reflecting that subclinical atrial fibrillation is only approximately in one-third of patients with stroke detected before an embolic event.^35^ This emphasizes the importance of detecting atrial fibrillation in the acute stage of ischemic stroke.

However, patients with strokes of an undetermined etiology also had a high SAP incidence. The reason for this may be that a high proportion of patients with stroke of undetermined etiology had a greater disability, unstable hemodynamic status, or even died during the acute phase; therefore, they might have a greater chance of being surveyed incompletely for stroke etiology. Another explanation is that many of these patients have several risk factors and comorbidities that directly influence the final TOAST classification, such as coexisting large-artery atherosclerosis and underlying conditions attributed to cardioembolism.

Consistent with previous studies, the NLR was higher in the SAP group, with the best cut-off of 2.5 in our study.^4, 7^ The NLR is a well-known marker of systemic inflammation and infection and also a predictor of prognosis in a variety of diseases, including malignancies and ischemic stroke.^36, 37^ More importantly, a high NLR has been in previous studies a predictor of SAP, probably due to poststroke immunosuppression and its association with stroke severity and infarct volume.^4, 7^

SAP prevention is important for the outcome of patients with acute stroke. Preventive antibiotics for SAP have been examined in previous studies and reported to be ineffective in reducing SAP incidence or improving patient outcomes.^38, 39^ Except for the early detection of SAP risk factors, including reversible and manageable END causes, pharmacological preventive methods exist. For example, angiotensin-converting enzyme inhibitors, which enhance the cough reflex, decrease the risk of pneumonia after stroke, especially in Asian populations, and are considered first-choice antihypertensive agents in older patients with repeated aspiration pneumonia according to the Japanese Society of Hypertension Guidelines.^40, 41^ Several studies conducted in Japan have shown the efficacy of cilostazol, a phosphodiesterase inhibitor, in preventing aspiration pneumonia. Cilostazol may increase dopamine and substance P concentrations in the brain, which are important for the swallowing reflex,^42^ and metoclopramide has been beneficial in patients fed via nasogastric tubes.^43^ In addition to pharmacological methods, maintaining oral hygiene, compensatory techniques, such as raising the bed head by at least 30°, the chin-down or chin-tuck maneuver for patients with dysphagia, and dietary modification, such as food thickening and pureeing, reduce the risk of aspiration pneumonia.^42^

This study has some limitations. First, prestroke independence was not formally recorded, which may have influenced the incidence of SAP. However, in the present study, we included only patients with a first-ever stroke, which may have lowered the impact of prior dependence, whereas patients with functional disabilities resulting from causes other than cerebrovascular disease could still be included. Second, the NIHSS is not routinely used as a bedside monitoring tool. We could not compare the NIHSS with the GCS in terms of their utility in defining END; however, the latter is a more broadly used and well-established scoring system in critical care, which may support its practicability. Finally, this was a single-center retrospective study with the inherent limitation of potential selection bias. In future research, additional external verification is required to arrive at definitive conclusions.

## CONCLUSIONS

The present study developed a simple predictive model for SAP, in which items are universally required for every patient with a stroke and can be easily accessed by multidisciplinary medical staff in the stroke team, not only physicians but also nurses. We also identified END after hospital arrival as an independent risk factor for SAP. Early identification of SAP risk factors and reversible END causes, followed by individualized prevention methods, may lower the incidence of SAP.

## ARTICLE INFORMATION

### Acknowledgments

The authors thank the personnel at the Health Examination Center and Department of Medical Education and Research of KVGH for providing information in response to inquiries and for assistance in data processing.

### NON-STANDARD ABBREVIATIONS AND ACRONYMS

AUC: area under the curve
CI: confidence interval
END: early neurological deterioration
GCS: Glasgow Coma Scale
ICD: International Classification of Diseases
NIHSS: National Institutes of Health Stroke Scale
NLR: neutrophil-to-lymphocyte ratio
OR: odds ratio
SAP: stroke-associated pneumonia
TOAST: Trial of Org 10172 Acute Stroke Treatment

